# Analysis of the Infection status and risk Factors of carbapenem-resistant Acinetobacter baumannii in children

**DOI:** 10.1101/2025.11.09.25339851

**Authors:** Liang Fan, Zhang Hui, Shang Yueyun, Ba Shuang Tianjin

**Affiliations:** Children’s Hospital (Tianjin University Children’s Hospital, rent of Critical Care Medicine, Tianjin Key Laboratory for the Prevention and Control of Infectious Diseases in Children, Tianjin 300074

**Keywords:** Children, Acinetobacter baumannil, Carbapenem antibiotics, Bacterial resistance, Risk factors

## Abstract

**Objective:** To analyze the clinical characteristics of Acinetobacter baumannii infection in hospitalized children in a tertiary children ‘s hospital in Tianjin and to explore the risk factors of infection. Methods: A retrospective study analyzed the data of Acinetobacter baumannii strains isolated from hospitalized children in the Department of Emergency and Critical Care Medicine at Tianjin Children’s Hospital from October 2019 to October 2024. Based on drug sensitivity results, they were divided into carbapenem-resistant group(CRAB) and carbapene n sensitive group (CSAB). The clinical characteristics, strain distribution, specimen sources, resistance profiles, and levels of inflammation or other markers of the two groups of children were compared and analyzed. Multivariate Logistic regression models were used to analyze the independent risk factors for CRAB infection. Results: A total of 112 children with AB infection were isolated in this study, including 66 cases of CSAB and 46 cases of CRAB, mainly from lower respiratory tract specimens (47.32%). The CRAB group was characterized by severe multidrug resistance (MDR) and pan-drug resistance (XDR). Multivariate Logistic regression analysis showed that invasive catheterization procedures, use of antibiotics within 3 months prior to admission (especially class >03 antibiotics), previous exposure to carbapenem antibiotics, and receiving immunosuppressive therapy were independent risk factors for CRAB infection in children. Conclusion: The situation of resistance to Acinetobacter baumannii is serious, and particular attention should be paid to the prevention and control of CRAB infection in the PICU.

Acinetobacter baumannii (CRAB) is a Gram-negative conditional pathogen. With the widespread use of broad-spectrum antibacterial drugs, especially carbapenems, in recent years, the number of carbapenem-resistant Acinetobacter has gradually increased, Multidrug resistance is widespread^[1]^. Infants and young children are more susceptible due to their lower immune function and often have multiple underlying diseases, and are more difficult to treat. This study retrospectively analyzed the drug resistance characteristics and risk factors of carbapenem resistant Acinetobacter baumannii (CRAB) infections in hospitalized children in the Department of Emergency and Critical Care Medicine of Tianjin Children’s Hospital over the past five years, providing evidence for the clinical prevention and treatment of such drug-resistant bacterial infections and related epidemiological studies.

## 1. Subjects and methods

### 1.1 Subjects

This study uses a single-center retrospective cohort study design. The subjects were 112 children admitted to the Department of Emergency and Critical Care Medicine at Tianjin Children’s Hospital from October 2019 to October 2024. Inclusion criteria: Hospitalized children aged younger than 18 years with symptoms of bacterial infection, confirmed as Acinetobacter baumannii by laboratory bacterial culture, and complete clinical electronic medical records. Exclusion criteria: Co-infection, presence of primary immunodeficiency underlying disease, etc. This study was reviewed and approved by the Ethics Committee of Tianjin Children’s Hospital and was exempted from informed consent, with the ethics number Lunshen W-2025-021.

### 1.2 Methods

#### 1.2.1 Clinical data

A retrospective analysis of Acinetobacter baumannii (AB) isolated from bacterial culture specimens in the Department of Emergency and Critical Care Medicine at Tianjin Children’s Hospital from October 2019 to October 2024. Repeated isolates from the same site of the same child were excluded, and only the first isolates were included. Collect medical records of the included cases,including age, gender, presence of underlying diseases, antibiotic use, history of invasive procedures, ICU admission,etc, to analyze and summarize the clinical characteristics of AB infection in children.

#### 1.2.2 Strain identification and drug sensitivity test

Strain identification was carried out using the fully automatic microbial VITEK 2 compact analyzer from Biomerieux, France. The antibiotic MIC values of the isolated Acinetobacter baumannii strains were determined using the microbroth dilution method. The drug sensitivity results were judged according to the 2017 American Society for Clinical and Laboratory Standards and were classified as sensitive (S), intermediate (I), and resistant (R). Carbapenem resistance is defined as resistance to either imipenem or meropenem and is divided into carbapenem resistance group (CRAB) and carbapenem non-resistance group (CSAB).

#### 1.2.3 Statistical analysis

Data analysis was performed using SPSS 29.0. Resistance rates were expressed as frequency (rate, %), and comparisons between groups were performed using x^2^ test, adjusted x^2^ test, or Fisher’s exact probability method. Measurement data conforming to the Normal distribution were expressed as mean ± standard deviation (x±s), and the independent sample t-test was used for comparisons between groups; Non-normally distributed data were expressed as median (interquartile range) [M (Q1,Q3)]and compared between groups using the Mann-Whitney U test. Multivariate logistic regression analysis was used to assess the risk factors for CRAB infection,and P < 0.05 was considered statistically significant.

## 2. Results

### 2.1 General Characteristics of Children with AB Infection

A total of 112 children with Acinetobacter baumannii infection were included in this study,among whom 66 cases (58.93%) were infected with CSAB, and 46 cases (41.07%) were infected with CRAB. The majority were male (63 cases, 56.25%), mainly in the 4-7 age group (37 cases, 33.04%). There was no statistically significant difference in gender composition and age distribution between the CSAB group and the CRAB group (P > 0.05). See Table 1 for details.

**Table 1.**
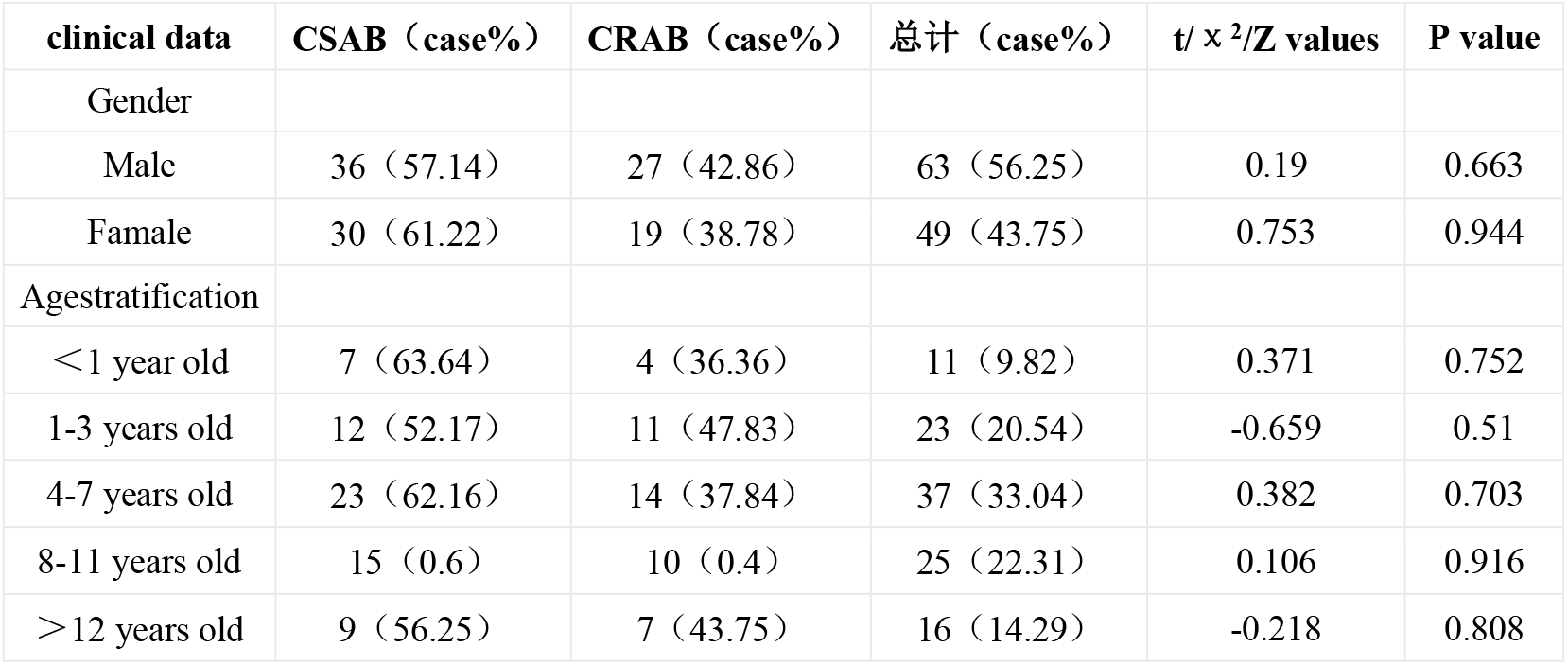
Clinical characteristics of gender and age distribution of AB infection in children in the case group.

| clinical data | CSAB (case%) | CRAB (case%) | 总计 (case%) | t/ $\chi^2$/Z values | P value |
| --- | --- | --- | --- | --- | --- |
| Gender |  |  |  |  |  |
| Male | 36 (57.14) | 27 (42.86) | 63 (56.25) | 0.19 | 0.663 |
| Famale | 30 (61.22) | 19 (38.78) | 49 (43.75) | 0.753 | 0.944 |
| Agestratification |  |  |  |  |  |
| <1 year old | 7 (63.64) | 4 (36.36) | 11 (9.82) | 0.371 | 0.752 |
| 1-3 years old | 12 (52.17) | 11 (47.83) | 23 (20.54) | -0.659 | 0.51 |
| 4-7 years old | 23 (62.16) | 14 (37.84) | 37 (33.04) | 0.382 | 0.703 |
| 8-11 years old | 15 (0.6) | 10 (0.4) | 25 (22.31) | 0.106 | 0.916 |
| >12 years old | 9 (56.25) | 7 (43.75) | 16 (14.29) | -0.218 | 0.808 |

### 2.2 Resistance

112 strains of Acinetobacter baumannii showed 100.00% resistance to both aztreonam and nitrofurantoin. Resistance rates to ampicillin, cefazolin, cefuroxime, and amoxicillin acid all exceeded 90.00%, with the CRAB group having 100.00% resistance rates to the above drugs, which may be associated with higher antibiotic exposure. In contrast, AB was relatively sensitive to quinolones such as ciprofloxacin, norfloxacin, and moxifloxacin, possibly due to the limited use of quinolones in pediatric clinical practice and fewer exposure opportunities in children,which led to a lower resistance rate. See Table 2 for more details.

**Table 2.** Resistance of 112 Acinetobacter baumannii strains [strains(%)]

| Antibiotics | CSAB | CSAB | CSAB | CRAB | CRAB | CRAB | Overall resistan-ce rate% |
| --- | --- | --- | --- | --- | --- | --- | --- |
| Antibiotics | Sensitive (e.g,%) | Intermedia ry (e.g,%) | Drug resistance (e.g,%) | Sensitive (e.g,%) | Intermedia ry (e.g,%) | Drug resistance (e.g,%) | Overall resistan-ce rate% |
| Ampicillin | 0 | 2 (3.03) | 64 (96.97) | 0 | 0 | 46 (100.00) | 98.21 |
| Amoxicillin sulbactam | 53 (80.30) | 0 | 13 (19.70) | 27 (58.70) | 3 (6.52) | 16 (34.78) | 25.89 |
| Piperacillin and tazobactam | 53 (80.30) | 0 | 13 (19.70) | 27 (58.70) | 3 (6.52) | 16 (34.78) | 25.89 |
| Amoxicillin clavulanate | 7 (10.61) | 2 (3.03) | 57 (86.36) | 0 | 0 | 46 (100) | 91.96 |
| Cefuroxime Sodium | 0 | 4 (6.06) | 62 (93.94) | 0 | 0 | 46 (100) | 96.43 |
| Ceftazidime | 40 (60.61) | 7 (10.61) | 19 (28.78) | 13 (28.26) | 9 (19.57) | 24 (52.17) | 38.39 |
| Cefotaxime | 40 (60.60) | 13 (19.70) | 13 (19.70) | 0 | 12 (26.09) | 34 (73.91) | 41.97 |
| Cefepime | 53 (80.30) | 0 | 13 (19.70) | 18 (39.13) | 2 (4.35) | 26 (56.52) | 34.82 |
| Ceftizoxime | 13 (19.70) | 0 | 53 (80.30) | 4 (8.70) | 2 (4.35) | 40 (86.95) | 83.04 |
| Norfloxacin | 66 (100) | 0 | 0 | 26 (56.52) | 2 (4.35) | 18 (39.13) | 16.07 |
| Ciprofloxacin | 66 (100) | 0 | 0 | 27 (58.70) | 2 (4.35) | 17 (36.95) | 15.18 |
| Moxifloxacin | 66 (100) | 0 | 0 | 36 (78.26) | 3 (6.52) | 7 (15.22) | 6.25 |

| Antibiotics | CSAB | CSAB | CSAB | CRAB | CRAB | CRAB | Overall resistance rate% |
| --- | --- | --- | --- | --- | --- | --- | --- |
| Antibiotics | Sensitive (e.g,%) | Intermediate (e.g,%) | Drug resistance (e.g,%) | Sensitive (e.g,%) | Intermediate (e.g,%) | Drug resistance (e.g,%) | Overall resistance rate% |
| Levofloxacin | 66 (100) | 0 | 0 | 36 (78.26) | 3 (6.52) | 7 (15.22) | 6.25 |
| SMZco | 54 (81.82) | 5 (7.58) | 7 (10.60) | 16 (34.78) | 2 (4.35) | 28 (60.87) | 31.25 |
| Tetracycline | 61 (92.42) | 5 (7.58) | 0 | 5 (10.87) | 1 (2.17) | 40 (86.96) | 35.71 |
| Tigecycline | 65 (98.48) | 1 (1.52) | 0 | 23 (50.00) | 2 (4.35) | 21 (45.65) | 18.75 |
| Ceftriaxone | 20 (30.30) | 21 (31.82) | 25 (37.88) | 2 (4.35) | 1 (2.17) | 43 (93.48) | 60.71 |
| Cefazolin | 0 | 4 (6.06) | 62 (93.94) | 0 | 0 | 46 (100) | 96.43 |
| Gentamicin | 58 (87.88) | 0 | 8 (12.12) | 16 (34.78) | 2 (4.35) | 28 (60.87) | 32.14 |
| Nitrofurantoin | 0 | 0 | 66 (100) | 0 | 0 | 46 (100) | 100 |
| Tobramycin | 65 (98.48) | 1 (1.52) | 0 | 32 (69.57) | 2 (4.35) | 12 (26.08) | 10.71 |
| Aztreonam | 0 | 0 | 66 (100) | 0 | 0 | 46 (100) | 100 |

### 2.3 Distribution of strain origin

The primary specimen source of 112 Acinetobacter baumannii strains in this study was the lower respiratory tract (sputum and bronchoalveolar lavage fluid), accounting for 47.32% (53/112);Followed by throat swabs (upper respiratory tract),which accounted for 21.43% (24/112). There was no statistically significant difference in the distribution of infection sources between the CSAB group and the CRAB group. See Table 3 for details.

**Table 3.** Distribution of sources of AB infection strains in the two groups of children (n, %).

| Sample Source | CSAB (n=66) | CRAB (n=46) | Total (%) | t/ $\chi^2$/Z values | P value |
| --- | --- | --- | --- | --- | --- |
| throat swab | 11 (16.67) | 13 (28.26) | 21.43 | 2.289<sup>a</sup> | 0.13 |
| punctate* | 5 (7.57) | 3 (6.52) | 7.14 | -<sup>b</sup> | 1.00 |
| Sputum and alveolar lavage fluid | 36 (54.54) | 17 (36.96) | 47.32 | 3.536<sup>a</sup> | 0.06 |
| urine | 8 (12.12) | 4 (8.69) | 10.71 | -<sup>b</sup> | 0.753 |
| Secretions* | 1 (1.52) | 3 (6.53) | 3.57 | -<sup>b</sup> | 0.301 |
| Pus | 5 (7.58) | 6 (13.04) | 9.82 | -<sup>b</sup> | 0.343 |

Note: a indicates Chi-Square Test, b indicates Fisher's exact test. \*: In this study, the puncture fluid mainly refers to the body fluids extracted through aseptic puncture, including cerebrospinal fluid, pleural and peritoneal effusion, joint fluid, etc. Secretions include specimens of soft tissue and wound secretions, as well as stoma discharge.

### 2.4 Analysis of risk factors for CRAB Infection

The proportions of children in the CRAB group who received mechanical ventilation and invasive catheterization, combined use of antibiotics within 3 months before admission, history of carbapenem antibiotic use, immunosuppressant treatment within 3 months, and stay in the intensive care unit (ICU) for more than 7 days were significantly higher than those in the CSAB group, and the differences were statistically significant (P<o.05). See Table 4 for details.

**Table 4.** Comparison of clinical data of two groups of children with AB infection [cases (%)]

| Classification of Situations | CSAB 组<br>(n=66) | CRAB 组<br>(n=46) | $\chi^2$ 值 | P 值 |
| --- | --- | --- | --- | --- |
| Combined underlying diseases | 39 (59) | 33 (71) | 1.9 | 0.618 |
| mechanical ventilation | 15 (23) | 19 (41) | 4.43 | 0.035 |
| Invasive catheterization | 21 (32) | 37 (80) | 25.13 | $<0.001$ |
| History of combined antibiotic use within 3 months | 18 (27) | 25 (54) | 8.31 | 0.004 |
| History of use of carbapenem antibiotics within 3 months | 14 (21) | 19 (41) | 5.27 | 0.022 |
| Had received immunosuppressive therapy within 3 months | 9 (14) | 26 (57) | 22.03 | $<0.001$ |
| Stay in ICU for at least 7 days | 15 (23) | 32 (70) | 24.83 | $<0.001$ |

### 2.5 Differences in inflammatory marker expression levels

There was no statistically significant difference in the median serum levels of inflammatory markers such as white blood cell count, neutrophil ratio, platelet count, C-reactive protein, procalcitonin, D-dimer, interleukin between The CRAB group and the CSAB group (P > 0.05), indicating that CRAB infection could not be determined based solely on the difference in infection indicators. See Table 5 for details.

**Table 5.**
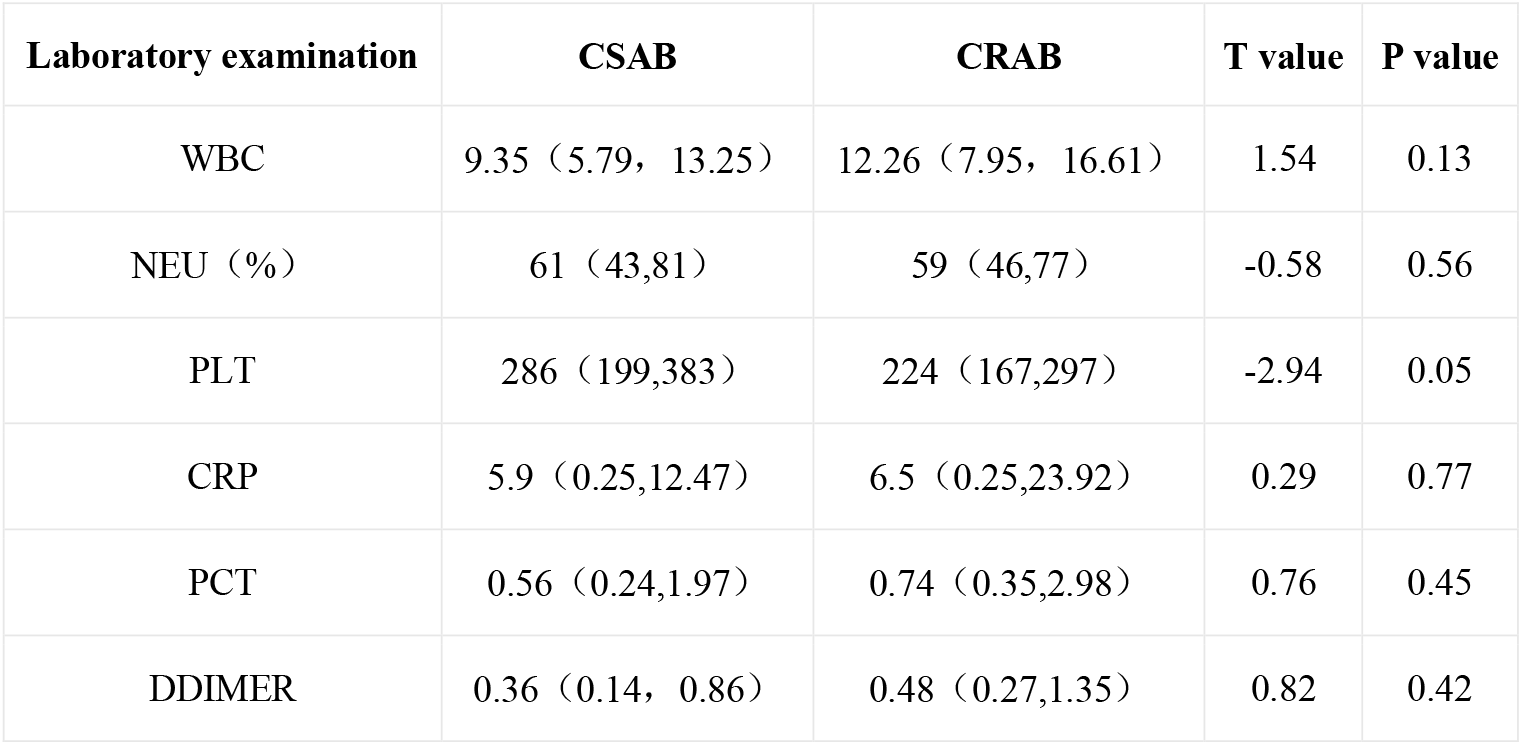
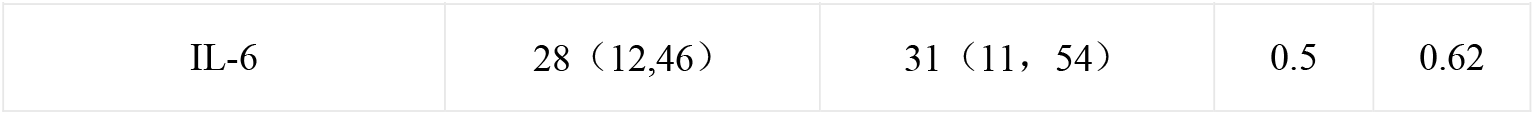
Comparison of Differences in Inflammatory Markers of AB infection between the two groups of children.

| Laboratory examination | CSAB | CRAB | T value | P value |
| --- | --- | --- | --- | --- |
| WBC | 9.35 (5.79, 13.25) | 12.26 (7.95, 16.61) | 1.54 | 0.13 |
| NEU (%) | 61 (43,81) | 59 (46,77) | -0.58 | 0.56 |
| PLT | 286 (199,383) | 224 (167,297) | -2.94 | 0.05 |
| CRP | 5.9 (0.25,12.47) | 6.5 (0.25,23.92) | 0.29 | 0.77 |
| PCT | 0.56 (0.24,1.97) | 0.74 (0.35,2.98) | 0.76 | 0.45 |
| DDIMER | 0.36 (0.14, 0.86) | 0.48 (0.27,1.35) | 0.82 | 0.42 |
| IL-6 | 28 (12,46) | 31 (11, 54) | 0.5 | 0.62 |

### 2.6 Independent risk factors for CRAB infection

Multivariate Logistic regression analysis showed that invasive catheterization procedures,a history of combined antibiotic use within 3 months prior to admission, exposure to carbapenem antibiotics, and immunosuppressive therapy within 3 months were all independent risk factors for CRAB infection (P<0.05). See Table 6 for details.

**Table 6.** Multivariate Logistic regression model analysis of Risk Factors for CRAB Infection in Children.

| Classification of Situations | Regression coefficient<br>($\beta$) | Standard Error<br>(SE) | Wald value<br>(Chi-square) | P value | OR | 95%CI |
| --- | --- | --- | --- | --- | --- | --- |
| Combined underlying diseases | -1.022 | 1.286 | 0.63 | 0.436 | 0.36 | 0.03-4.65 |
| mechanical ventilation | 0.65 | 0.48 | 1.83 | 0.173 | 1.93 | 0.75-4.9 |
| Invasive catheterization | 2.1 | 0.5 | 17.64 | $< 0.001$ | 8.16 | 3.06-21.76 |
| History of combined antibiotic use within 3 months | 1.35 | 0.38 | 7.9 | 0.005 | 3.86 | 1.51-9.85 |
| History of use of carbapenem antibiotics within 3 months | 0.9 | 0.52 | 3 | 0.033 | 2.46 | 0.89-6.8 |
| Had received immunosuppressive therapy within 3 months | 1.85 | 0.55 | 11.29 | 0.001 | 6.36 | 2.17-18.63 |
| Stay in ICU for at least 7 days | 0.4 | 0.3 | 1.78 | 0.177 | 1.49 | 0.84-2.68 |

### 2.7 Independent risk factor analysis for invasive CRAB infection

Literature reports that AB is often difficult to distinguish between colonization and infection. The key points for differentiation are as follows: l. Lack of clear infection-related symptoms and signs; 2. The child is generally in good condition, with no or only minor underlying diseases;3. Inflammatory markers are normal or slightly elevated without dynamic elevation; 4. The culture is a mixture of multiple bacteria, and non-dominant bacteria, and the quantitative culture quantity is below the threshold; 5.No medication was administered against Acinetobacter baumannii, and the child’s condition did not deteriorate or even improve. Only medication was administered against Acinetobacter baumannii, but there was no clinical improvement. Based on the above criteria, 17 of the 46 CRAB cases in this study were identified as non-invasive infections, accounting for 36.96%, and 29 were identified as invasive infections, accounting for 63.04%. Multivariate Logistic regression analysis show that coexisting underlying diseases and staying in the ICU for more than 7 days were independent risk factors for invasive CRAB infection in children (P<0.05), as shown in Table 7.

**Table 7.** Multivariate Logistic regression model analysis of Risk factors for Invasive CRAB Infection in children.

| Classification of Situations | Regression<br>coefficient<br>($\beta$) | Standard<br>Error<br>(SE) | Wald value<br>(Chi-square) | P value | OR | 95%CI |
| --- | --- | --- | --- | --- | --- | --- |
| Combined underlying diseases | 3.58 | 1.12 | 10.27 | 0.001 | 35.89 | 4.33-297.55 |
| mechanical ventilation | -0.494 | 0.865 | 0.33 | 0.563 | 0.61 | 0.11-3.26 |
| History of combined antibiotic use within 3 months | -0.545 | 0.798 | 0.47 | 0.491 | 0.58 | 0.12-2.74 |
| History of use of carbapenem antibiotics within 3 months | 1.015 | 0.947 | 1.36 | 0.243 | 3.02 | 0.47-19.23 |
| Had received immunosuppressive therapy within 3 months | 0.191 | 0.802 | 0.06 | 0.81 | 1.21 | 0.25-5.8 |
| Stay in ICU for at least 7 days | 2.97 | 0.92 | 10.32 | 0.001 | 19.5 | 3.57-106.60 |

## 3. Discussion

Acinetobacter baumannii is one of the significant pathogens causing nosocomial infections in China^[2]^. Its resistance rate to carbapenem antibiotics is close to 65%^[1]^, and it has been classified as a “critical priority” pathogen by the World Health Organization^[3]^. The short-term mortality rate of patients with hospital-acquired CRAB infection is 1.3 to 6.9 times higher than that of carbapenem-sensitive strains^[4]^.Children have underdeveloped immune systems and are susceptible to nosocomial infections. In this study, most of the children had a history of medical visits and antibiotic use before infection. The 112 infected children covers all age groups,but there were no significant differences in gender composition and age distribution between the CSAB group and the CRAB group.

In recent years, the clinical isolation rate of multidrug-resistant Acinetobacter baumannii has increased significantly. The mechanism of resistance is complex, involving the production of β-lactamases, overexpression of efflux pumps, mutations in antibiotic targets, and changes in the permeability of outer membrane proteins^[5]^. Acquired carbapenemase genes were detected in 91% of CRAB isolates, mainly mediated by Class B (metal-β-lactamases) and Class D (such as OxA-type carbapenemases)^[6‵ 7]^. Among them,OXA-23 type D is the most significant determinant of resistance in CRAB, accounting for up to 99%, making it the dominant genotype of CRAB in China^[8]^. Studies have found that Acinetobacter baumannii is prone to causing nosocomial infections and is a multidrug-resistant strain. It is difficult to treat and has a high fatality rate. Once AB is resistant to carbapenems^[9]^, it often develops cross-resistance to multiple other antibiotics (such as β-lactam, quinolones, etc.) at the same time^[10]^. This is consistent with the results of this study showing severe multidrug resistance (MDR) and pan-drug resistance (XDR) in the CRAB group.

The study showed that AB strains were mainly isolated from lower respiratory tract specimens, consistent with the highest detection rate of CRAB in alveolar lavage fluid, as reported by the National Bacterial Resistance Surveillance Network (CARSS) in 2023. Notably, the infection rate of CRAB rose significantly during the novel coronavirus pandemic^[11]^. This study confirmed that mechanical ventilation, invasive catheterization, antibiotic use three months before admission, carbapenem exposure, and immunosuppressant therapy were all independent risk factors for CRAB infection, consistent with the conclusions of multiple studies at home and abroad^[12‵ 13‵ 14‵ 15]^. The study shows that the pathogenic mechanism of CRAB is closely related to its strong biofilm-forming ability^[12‵ 14]^, and AB isolates can form stable biofilms on the surface of medical devices such as central venous catheters and tracheal intubations^[16]^. Children in the pediatric intensive care unit (PICU) have a significantly increased risk of CRAB infection due to weakened immune function, long hospital stays, and frequent invasive procedures. Multiple studies have pointed out that pre-infection antibiotic exposure can disrupt the host’s normal microbiota barrier^[17]^, stimulate genetic variations in pathogenic bacteria, promote the generation and horizontal transfer of resistance genes, and accelerate the spread of resistance^[18]^. Carbapenem antibiotics are currently the commonly used empirical treatment for severe Gram-negative bacterial infections, but the irrational use of carbapenem drugs, such as below the minimum inhibitory concentration, can affect the expression of penicillin-binding protein coding genes in Acinetobacter baumannii, thereby leading to drug resistance^[19]^.

Clinical treatment of CRAB infection faces many challenges. It is difficult to determine whether it is the direct cause of death based on microbiological results alone^[18]^. Colonization and infection need to be differentiated in combination with clinical features and treatment outcomes. In this study, the proportion of children with invasive CRAB infection who had underlying diseases, ICU admission, and a history of carbapenem antibiotic exposure was significantly higher than that in the colonization group. Among them, underlying diseases and ICU admission were independent risk factors for invasive CRAB infection. This result suggests that the risk of CRAB infection in critically ill children with underlying diseases should be closely vigilant in clinical practice. Improvement of diagnostic accuracy. Studies show there is no “standard antibacterial regimen” for CRAB infections at present^[20]^. In this study, although the resistance rates of quinolones (such as ciloxacin and moxifloxacin), tigecycline, and tetracyclines were relatively low, due to the special limitations of pediatric use, the options for clinical anti-infection treatment are extremely limited. Currently, at least two drug combinations are mostly recommended in clinical practice, and monotherapy should be avoided as much as possible^[21]^.

In summary, when dealing with critically ill children with Acinetobacter baumannii infection, clinicians should attach great importance to the prevention and control of CRAB infection in the PICU, select antibacterial drugs as accurately and properly as possible based on drug sensitivity. results, strictly control the indications for the use of carbapenem antibiotics, avoid empirical overuse, and strengthen disinfection, isolation, and hospital infection control. To curb the generation and spread of CRAB as much as possible. The limitations of this study lie in the Single-center analysis, the small number of specimens from clean or non-clean open isolates of Acinetobacter baumannii, and further research with larger sample sizes is needed later.

## Data Availability

All relevant data are within the manuscript and its Supporting Information files

